# Rapid and accurate identification of COVID-19 infection through machine learning based on clinical available blood test results

**DOI:** 10.1101/2020.04.02.20051136

**Authors:** Jiangpeng Wu, Pengyi Zhang, Liting Zhang, Wenbo Meng, Junfeng Li, Chongxiang Tong, Yonghong Li, Jing Cai, Zengwei Yang, Jinhong Zhu, Meie Zhao, Huirong Huang, Xiaodong Xie, Shuyan Li

## Abstract

Since the sudden outbreak of coronavirus disease 2019 (COVID-19), it has rapidly evolved into a momentous global health concern. Due to the lack of constructive information on the pathogenesis of COVID-19 and specific treatment, it highlights the importance of early diagnosis and timely treatment. In this study, 11 key blood indices were extracted through random forest algorithm to build the final assistant discrimination tool from 49 clinical available blood test data which were derived by commercial blood test equipments. The method presented robust outcome to accurately identify COVID-19 from a variety of suspected patients with similar CT information or similar symptoms, with accuracy of 0.9795 and 0.9697 for the cross-validation set and test set, respectively. The tool also demonstrated its outstanding performance on an external validation set that was completely independent of the modeling process, with sensitivity, specificity, and overall accuracy of 0.9512, 0.9697, and 0.9595, respectively. Besides, 24 samples from overseas infected patients with COVID-19 were used to make an in-depth clinical assessment with accuracy of 0.9167. After multiple verification, the reliability and repeatability of the tool has been fully evaluated, and it has the potential to develop into an emerging technology to identify COVID-19 and lower the burden of global public health. The proposed tool is well-suited to carry out preliminary assessment of suspected patients and help them to get timely treatment and quarantine suggestion. The assistant tool is now available online at http://lishuyan.lzu.edu.cn/COVID2019_2/.

**Funding:** This work was supported by the Fundamental Research Funds for the Central Universities (lzujbky-2020-sp11) and the Gansu Provincial COVID-19 Science and Technology Major Project, China.

## Introduction

Coronavirus disease 2019 (COVID-19) ^1^ can induce emerging acute respiratory illness by severe acute respiratory syndrome coronavirus 2 (SARS-CoV-2) which is classified by International Committee on Taxonomy of Viruses (ICTV).^2^ This enveloped RNA virus is the seventh member of the coronavirus family to infect humans belonging to lineage B of the genus betacoronavirus through deep sequencing and phylogenetic analysis.^3^ The epidemic pneumonia has spread quickly and become a global public health concern.^4^ After making careful assessment for COVID-19, the epidemic is classified as a pandemic due to the alarming spread of the infection and the severity of the disease in March 11, 2020.^5^ A large amount of practice and research show that the common clinical symptoms of the pneumonia are fever, cough, myalgia or fatigue.^6^ And the characteristics of chest radiography or computed tomography (CT) are multiple mottling or ground-glass opacity.^7^

According to these valuable experience and achievements, the seventh edition of a new coronavirus pneumonia diagnosis and treatment plan has jointly updated by two Chinese departments. The official guidelines emphasize that the use of quantitative real-time PCR (qRT-PCR) assays for the detection of viral nucleic acid to diagnose the infected patients is still one of the main reference standards.^8^ Although RNA of this coronavirus is the prime marker that can be detected in the early stage of infection, the high requirements of its testing technology and condition limit its positive rate to as about 30-50%.^9^ This situation may imply that many patients with COVID-19 cannot be identified in time.^10^ The situation may delay the isolation and treatment of infected patients and accelerate the spread of the epidemic due to the characteristic of human-to-human transmission.^11^ Apart from this, some objective factors, such as complicated operation, time-consuming, easy to pollute, high cost and so on, further hinder the technology from exerting its due advantages on emergency prevention and control conditions.^12^ Many clinicians strongly recommended the use of CT imaging in severe epidemic areas, however, the tardy method would present obvious abnormalities when the lung produced inflammatory or the tissue generated lesions.^13^ At present, the combined detection of immunoglobulin M (IgM) and immunoglobulin G (IgG) antibody provides a fast and simple screening method, but the two substances are impossible to be detected in the window period of about two weeks.^14,15^ So this method can only be used as an effective supplement and auxiliary diagnosis of nucleic acid detection.^16^

Until now, the quick diagnosis of COVID-19 patients in a large scale is still a major problem for global health concern.^17^ Except for the laboratory-confirmed COVID-19 patients, there are a large number of suspected patients and close contacts who need to be confirmed whether they are infected or not.^18^ In this case, it is difficult to screen and identify the cases during the incubation period or subclinical infections, they have great potential to become super spreaders.^19^ Although timely isolation measures can effectively cut down the spread of the virus, early diagnosis can enable patients to get more active treatment at the initial stage of presentation.^20^ Therefore, the early identification and timely management of COVID-19 patients is of great importance due to no specific treatment.^21^

The most common laboratory findings, including hematological and biochemical parameters, play a prominent part in the initial screening for COVID-19.^22^ In the present work, we use machine learning algorithm to process these valuable parameters in order to realize the progress of accurate and rapid identification for the emerging pneumonia. The proposed assistant discrimination tool is more convenient and practical than nucleic acid detection and antibody detection, which can accelerate the recognition of suspected patients and thus reduce the risk of transmission. Moreover, the deep relationship between these important reference targets and COVID-19 could help to monitor the progress of the disease and to guide useful strategies for clinicians. Finally, we aim to design a simple and effective pre-warning system to help proactively initial screening for suspected patients which will contribute to the outbreak control and public health.

## Materials and Methods

### Source of materials

In this study, a total of 253 samples from 169 suspected patients were collected from multiple sources, including Lanzhou Pulmonary Hospital, the First Hospital of Lanzhou University, Lanzhou University Second Hospital, the First People’s Hospital of Lanzhou City, and Gansu Provincial Hospital. Data from routine laboratory assessments with 49 parameters for each sample included 24 routine hematological parameters and 25 biochemical parameters. General distribution information about the datasets can be listed in Table 1 and the detailed information about the patients, including sex, age, and 49 parameters are listed in Table S1. 105 consecutive samples from 27 patients with confirmed COVID-19 admitted to Lanzhou Pulmonary Hospital in Gansu Province were considered as positive samples and randomly divided into training set, test set, and external validation set. The virus nucleic acid assays (qRT-PCR) of throat swab and sputum samples were detected for all these patients to make a definite diagnosis. The diagnostic evidence for COVID-19 is based on World Health Organization interim guidance. Except for the COVID-19 samples, the remaining samples, which were collected from patients with similar symptoms or similar radiologic characteristic with COVID-19, including patients with common pneumonia, tuberculosis and lung cancer, were treated as negative samples. These patients were diagnosed by at least two professional experts based on comprehensive examination results. Among them, pneumonia samples were collected from multiple sources. The pneumonia samples in training set and test set were collected from the First Hospital of Lanzhou University, the First People’s Hospital of Lanzhou City, and Gansu Provincial Hospital. The pneumonia samples in external validation set were from Lanzhou University Second Hospital. The tuberculosis and lung cancer samples were also collected from Lanzhou University Second Hospital. Clinical data collection from patients was approved by Ethics Committee of the First Hospital of Lanzhou University which is the leading department for this project.

**Table 1.** Distribution and baseline characteristics of the whole data

|  | Training set | Test set | External validation set | Total number | Median age (IQR) | Male/Female |
| --- | --- | --- | --- | --- | --- | --- |
| COVID-19 | 50 (12)* | 15 (5)* | 40 (10)* | 105 (27)* | 47 (33-64) | 13/14 |
| Common pneumonia | 76 | 8 | 14 (8)* | 98 (92)* | 63 (47-75) | 44/48 |
| Tuberculosis | 10 | 5 | 10 | 25 | 54 (33-68) | 15/10 |
| Lung cancer | 10 | 5 | 10 | 25 | 61 (54-69) | 19/6 |

\*Note: The number inside the bracket is the total number of patients where all the serial samples were collected

### Machine learning method

Random forest algorithm (RF) is a promising and well-known classifier with praiseworthy characteristics that can use multiple trees to train and predict samples, which has received extensive attention in the fields of chemometrics and bioinformatics.^23,24^ The algorithm in this study was used to establish the final assistant discrimination tool to proactively seek out patients with COVID-19. The whole process could be divided into three stepwise stages. In the first stage, all the parameters were fully employed to build the model in order to evaluate the importance of each parameter. In the second stage, while the top-ranking parameters were increased one by one, different models were built under the adjustment of RF parameters including the tree number (ntree) and the number of randomly selected features to split at each node (mtry). The adjustment range of ntree ranged from 500 to 1500 with a step of 100; Once the value of ntree was fixed, mtry spanned from 2 to 15 via a step of 1. In the last stage, based on the principle that the minimum number of top-ranking parameters could have comparable performance with the whole parameters, the appropriate number of parameters, ntree, and mtry were selected respectively. The study was executed on R package randomForest v4·6-7.

### Validation method

In this work, several different validation methods are performed to ensure the reliability and reproducibility of this COVID-19 assistant identification tool.

Firstly, the internal 10-fold cross-validation which was performed on the training set was a persuasive and universal method to obtain robust performance. Secondly, the test set is totally different from the training set just to verify the discriminating ability of the tool. In addition, all modeling processes were accomplished in the training set based on 10-fold cross-validation and had no connection to the test set. Finally, after building the model with the selected optimal parameters, the external validation set was used to reconfirm the accuracy and repeatability of the tool.

There are some popular metrics to comprehensively assess the performance of the assistant tool including sensitivity (Sens), specificity (Spec), total accuracy (ACC), Matthews correlation coefficient (MCC), and area under the curve (AUC).

## Results and discussion

### Modeling results

It was highly important to select appropriate number of parameters for the foundation of the final discrimination tool with the advantages of convenient operation and accurate diagnosis. Based on the minimum number of top-ranking parameters without compromising performance, 11 laboratory findings were carefully selected as the final input indicators as shown in Figure 1. The final model revealed good prediction performance on the training set with ACC, MCC, and related AUC values of 0·9795, 0·9594, and 1·000, respectively. These important metrics implied that the tool integrated with 11 top-ranking parameters had the significant ability to discriminate patients with COVID-19. At the same time, the improvement of sensitivity (0·9750) and specificity (0·9909) heralded that this method had great potential as a practical clinical tool for large-scale initial screening. Although 34 parameters seemed like a sensible choice, the redundant ones didn’t play an irreplaceable role in sharply improving the performance, which might make the model over-fitting and prolong the diagnosis process. So the selected parameters were of extraordinary benefit to build a cost-effective and rapid assistant tool which had commensurate ability compared with the whole parameters.

**Figure 1:**
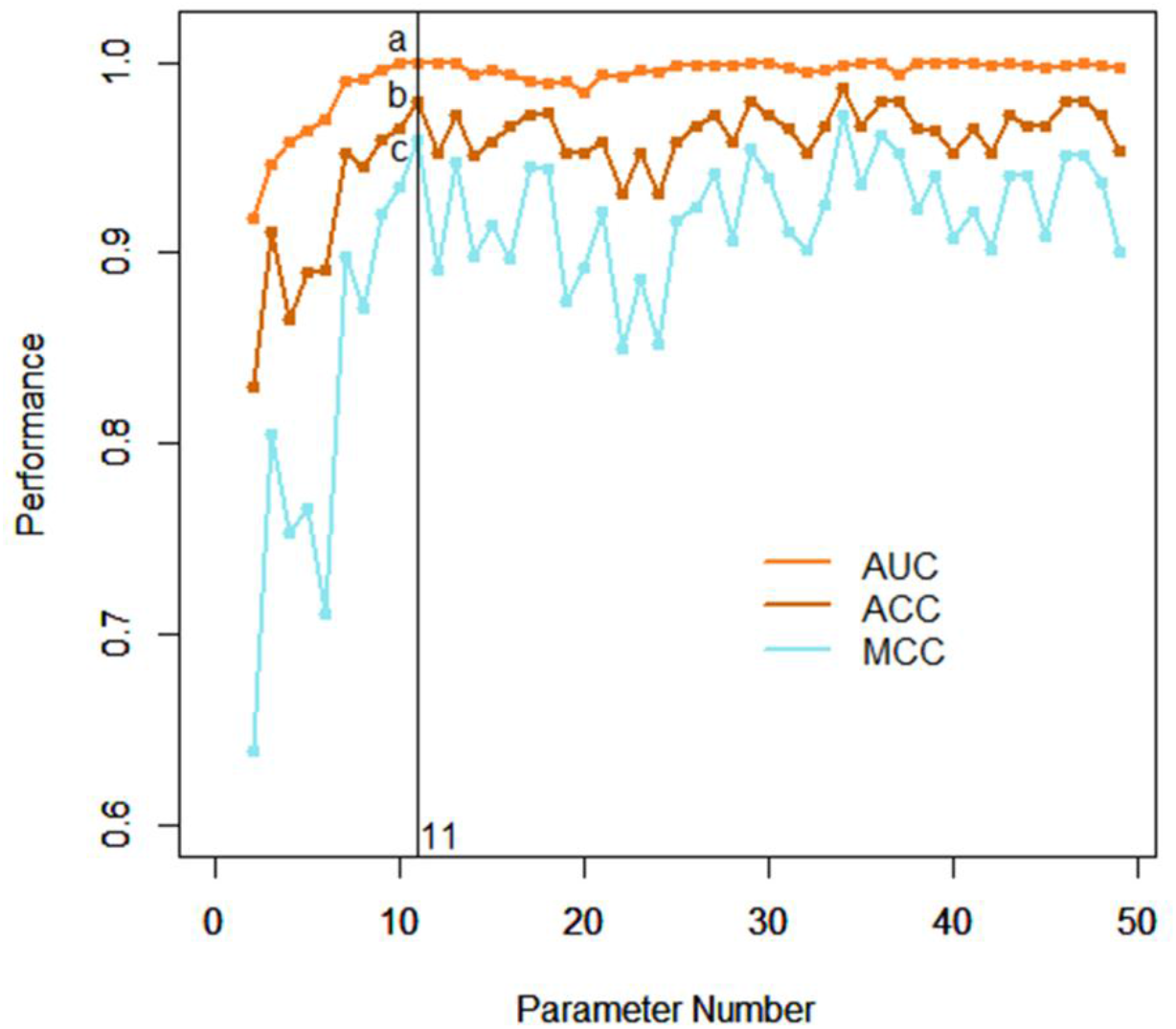
Performance of the various models. With the gradual increase of top-ranking parameters, the performance of different models is obtained based on the 10-fold cross-validation results in the training set (a=1·000; b=0·9795; c=0·9594).

The above data raised the hope that the tool, which combined machine learning algorithm and laboratory parameters, would play a prominent part in screening patients with COVID-19. The test set was completely independent of the entire modeling process and was adopted to prove the stability and reliability of this method again. As shown in Figure 2, the performance of the method in the test set was consistent with the training set with an AUC of 0·9926, achieving a sensitivity of 1.0000 and a specificity of 0·9444. It confirmed that the discrimination tool had the ability to deal with a large number of suspected patients with COVID-19, which might greatly simplify the laboratory blood test process and provide timely treatment for the confirmed patients.

**Figure 2:**
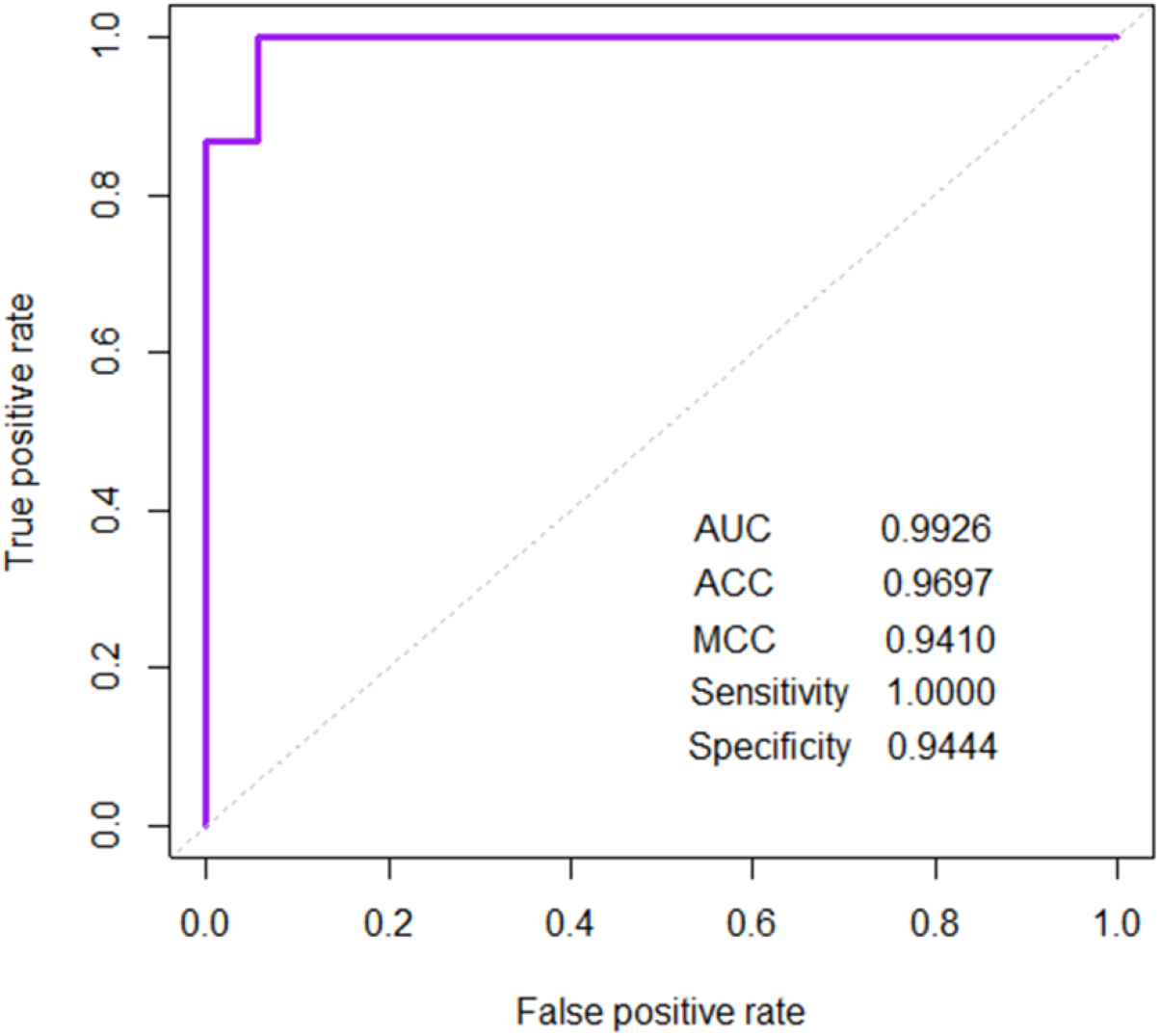
The ROC curve and discrimination performance for the test set. After the specific top-ranking parameter are selected, the performance of the built model on the test set is measured by some metrics.

### Application of this identification tool on external validation set

Under the guidance of the outstanding performance, the final model was built based on the sophisticated combination of 11 selected parameters. According to the performance requirements, the value of cutoff was adjusted to 0·43. The external validation set with 11 laboratory parameters also didn’t participate in any modeling process and was only used to reconsider the reliability of the model, which could provide the factual foundation for future clinical applications. Among these samples, 39 of 40 samples with COVID-19, 13 of 14 samples with common pneumonia, 10 of 10 samples with tuberculosis, and 9 of 10 samples with lung cancer were accurately identified. Overall, the sensitivity reached 39/(39+1+1) = 0·9512, the specificity reached 32/(32+1) = 0·9697, and the total accuracy reached 0·9595. These results indicated that the assistant tool might have a high resolution on distinguishing patients with COVID-19 from the complex disease condition. It suggested that the technology had great potential to develop into a powerful assistant tool for large-scale detection of the highly infectious disease.

### Key blood indices that were highly related with COVID-19

Only the integration of 11 laboratory parameters could achieve high-level performance to accurately identify COVID-19. The reason was not only the powerful ability of machine learning algorithm in medicine, but also the inextricable connection between the selected parameters and the epidemic pneumonia. Once the virus invaded the body, the composition of the blood would change. The abnormalities sometimes cannot be observed directly from these routine laboratory blood tests, but the parameters could play a material impact on identifying the disease after processed by machine learning. Subsequently the laboratory findings were analyzed from the statistical perspective and the detailed information of the findings was listed in Table 2S, including the abbreviations and reference ranges. Except for glucose (GLU) and magnesium (Mg), almost all the parameters manifested the significant differences between COVID-19 and general pneumonia in Figure 3. It also indicated the parameters were highly correlated to the infection of SARS-CoV-2 and the powerful ability of the tool to effectively distinguish patients with COVID-19 and patients with pneumonia. In fact, many abnormal changes of laboratory parameters had been widely reported for important clinical references, including total bilirubin (TBIL), GLU, creatinine (CREA), lactate dehydrogenase (LDH), creatine kinase isoenzyme (CK-MB), and kalium (K).^22,25,26^ For example, the descriptive study of 99 cases with COVID-19 in Wuhan showed that 51 (52%) patients with the elevated level of GLU and 75 (76%) patients with the elevated level of LDH, which was consistent with our results.^27^ The remaining parameters, including total protein (TP), calcium (Ca), magnesium (Mg), platelet distribution width (PDW), and basophil (BA#), were rarely received extensive attention due to the lack of information on the clinical characteristics of affected patient. However, they played an irreplaceable part in the random forest algorithm and had great potential for diagnostic markers with future clinical practice. Indeed, the existence of differences was not a decisive factor to select critical parameters for machine learning algorithms, it was helpful to promote the tool to quickly identify COVID-19 infection and provide a sufficient consciousness for dynamically monitoring the process of the disease.

**Figure 3:**
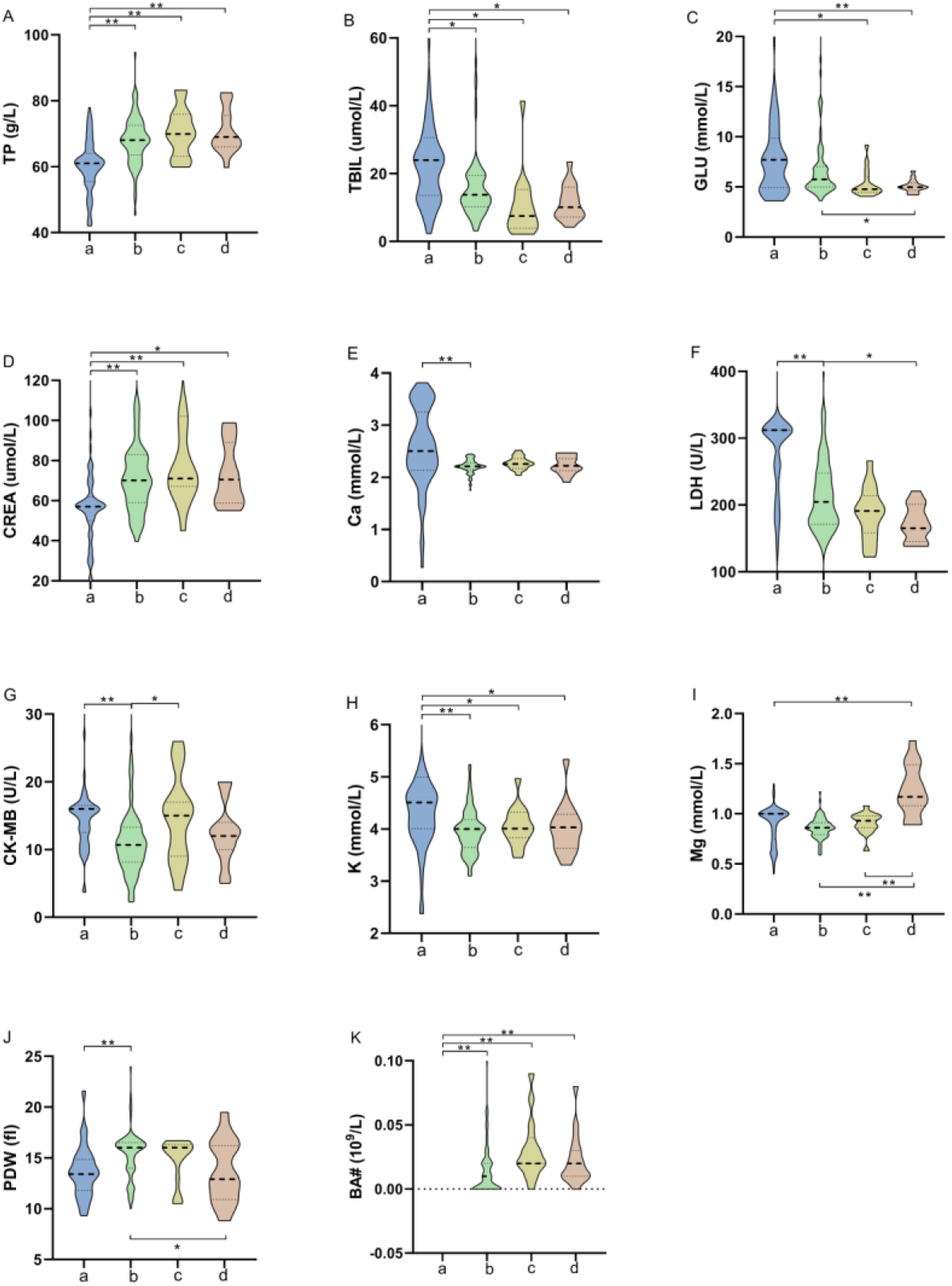
Differential expressed parameters in different patients’ groups. The statistical variations of 11 parameters are analyzed from different groups including group of COVID-19 (a), group of common pneumonia (b), group of tuberculosis (c), and group of lung cancer (d). ** represents p<0·0001 except for CLU (** p=0·0076), CREA (** p=0·0022 for a and b, ** p=0·0002 for a and c), LDH (** p=0·0047), and BA# (** p=0·0004 for a and b). TBIL (* p=0·018 for a and b, p=0·021 for a and c, p=0·029 for a and d), GLU (* p=0·016 for a and c, p=0·037 for b and d), CREA (* p=0·015), LDH (* p=0·038), CK-MB (* p=0·025), K (* p=0·036 for a and c, p=0·030 for a and d), PDW (* p=0·020).

### Complementarity with other analytical techniques

Currently, suspected patients were recommended to use nucleic acid assay to effectively control the spread of the epidemic pneumonia. The complexity of the operation and the fragility of the specimens make it unsuitable for large-scale initial screening. The time-consuming etiology testing might delay the timely isolation and treatment for COVID-19 patients. And a study reported that the positive rate of the testing was only 30-50% in clinical application.^9,28^ The sequencing technology of the viral had elevated the accuracy of detection results, but it needed high-quality laboratory requirement which could only be provided for supplementary evidence. Rapid antibody detection often lagged behind the nucleic acid detection due to the presence of the window period.^14^ After the virus invaded, the dynamic balance was often broken and the composition was changed in the blood. The timely responses might be used for the identification of the disease as long as they could be captured by machine learning algorithms. And the assistant tool was not subject to complex operation and prolonged waiting because the conventional laboratory blood tests were the mature technology that could be fully automated. It was convenient to operate the large-scale initial screening and help the confirmed patients provide appropriate treatment.

### Web server of the assistant tool

Finally, a user-friendly website service was built to experience and test the assistant tool online at http://lishuyan.lzu.edu.cn/COVID2019_2/. The 11 selected parameters and more detailed information were listed in Figure 4. The users only needed to input the value of specified parameters in the corresponding text box based on the instructions of the interface, and then clicked the “Submit” button. After calculation and analysis of the tool, the results page would display whether this sample was classified as a COVID-19 patient with the probability of prediction. It was vital to check that the units of laboratory parameters were consistent with those on the main page. Importantly, the tool could only be applied to test this method at present, and it is insufficient to evolve the definite evidence of clinical diagnosis for directly discriminating the COVID-19 patients.

**Figure 4:**
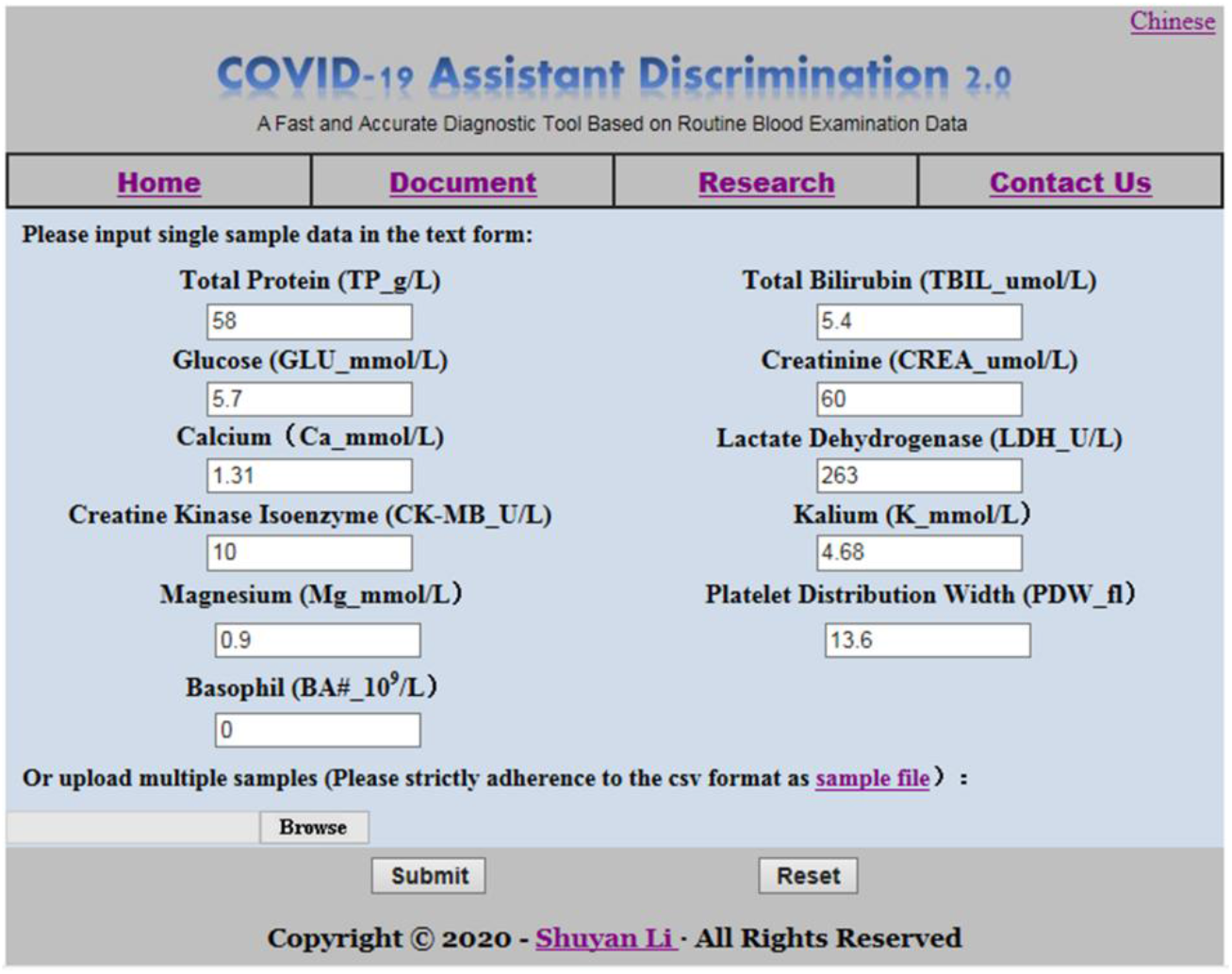
The main page of the COVID-19 assistant discrimination web server. Users only need to input 11 corresponding parameters according to the requirements of the above interface; the web server can display the result whether the sample is COVID-19.

### Further clinical evaluation on imported cases from outside of China

After the construction of the assistant discrimination tool was completed, there are some confirmed COVID-19 cases that were imported from outside of China and were received in Gansu Province. In order to make an in-depth assessment for the built tool, 24 samples of 14 patients from Lanzhou Pulmonary Hospital were collected when first performed the routine laboratory blood tests, including 2 routine hematological parameters and 9 biochemical parameters. The prediction results were shown in Figure 5. 22 of 24 samples were accurately identified through using our designed web server with an accuracy rate of 0·9 167. Changes for related blood components could not be captured by the tool due to the early stages of virus infection, the patients with mild symptoms were not accurately diagnosed. However, the high-precision performance has greatly encouraged the confidence to make more adequately preparation for further clinical practice.

**Figure 5:**
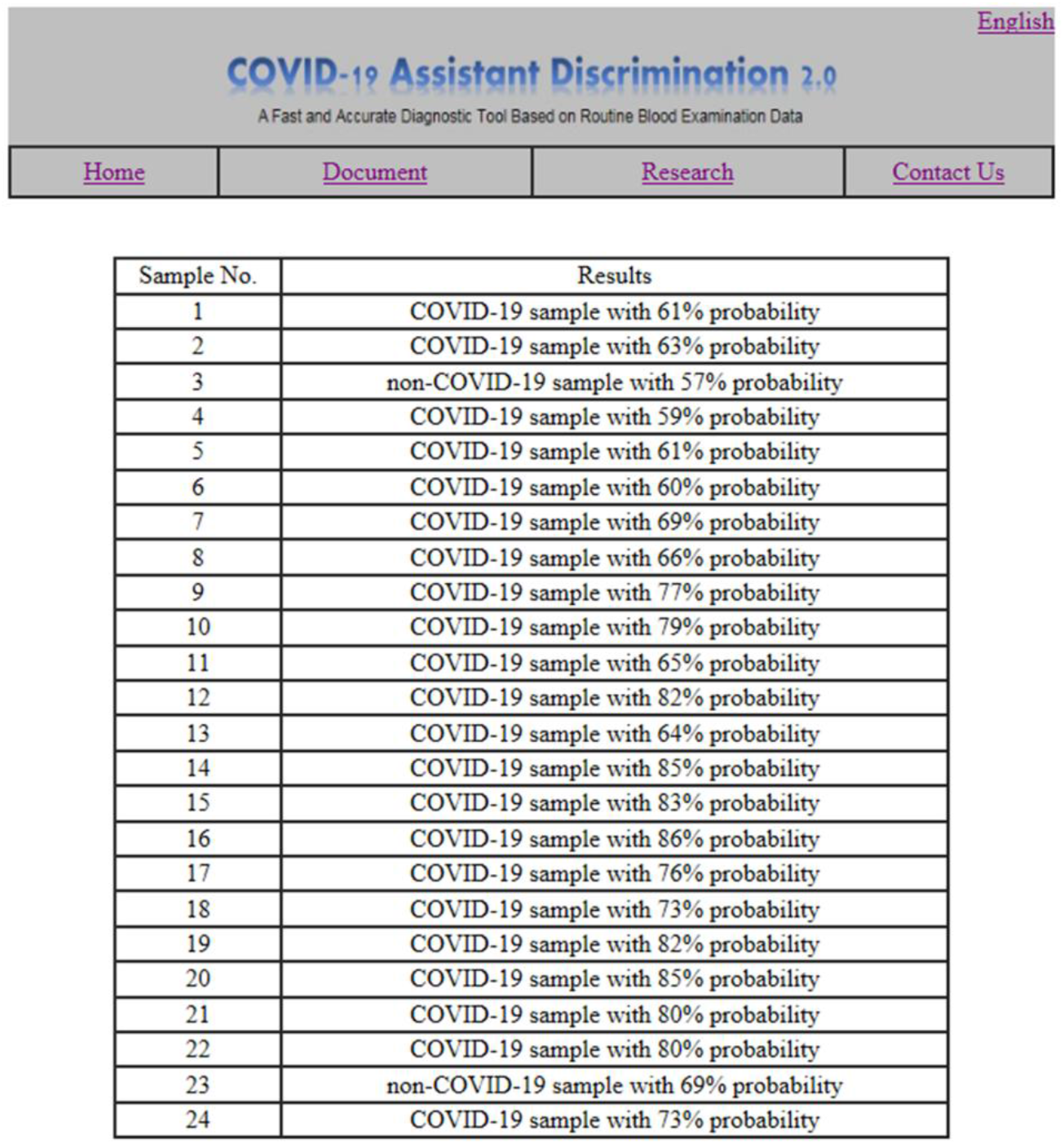
Prediction results of laboratory confirmed patients with COVID-19 overseas. 24 confirmed COVID-19 cases that were imported from outside of China are tested by the assistant discrimination tool.

### Application domain of this tool

This method is built on data of suspected patients which presented similar CT information or similar symptoms with COVID-19 like fever, cough, myalgia or fatigue, which are followed the first triage results in hospital before the nucleic acid assay. This tool has been validated many times with good performance on these suspected patients and we have faith on its proper application on further tests to identify COVID-19 cases from other suspected cases.

However, its application on identification COVID-19 cases with atypical symptoms is not confirmed since these cases were hard to collect in the current situation. Anyway, we still believe that if more laboratory parameters are collected, machine learning algorithms may extract more valuable candidate markers to identify COVID-19 infection, even from the whole population, whether with or without typical symptoms.

## Conclusion

In conclusion, the assistant tool was built with 11 top-ranking clinical available blood indices that were extracted from the random forest algorithm. This result may capture the inherent patterns of these routine parameters to make routine blood tests play an influential role on the value of alarm diseases. After several evaluations, it is of extraordinary benefit to be exploited for large-scale initial screening on suspected patients of COVID-19 to minimize the risk of the virus, especially for confirmed patients taken the timely isolation and treatment, which is currently known the most effective response. Although the proposed tool needs to be tested by more clinical practice, the method provides some new insights into for the rapid diagnosis of COIVD-19 infection to cope with severe situations due to terrible characteristic of the human-to-human transmission.

## Data Availability

The data used to support the findings of this study are available from the corresponding author upon request.

